# Correlation between post-vaccination titres of combined IgG, IgA, and IgM anti-Spike antibodies and protection against breakthrough SARS-CoV-2 infection: a population-based longitudinal study (COVIDENCE UK)

**DOI:** 10.1101/2022.02.11.22270667

**Authors:** Giulia Vivaldi, David A Jolliffe, Sian Faustini, Hayley Holt, Natalia Perdek, Mohammad Talaei, Florence Tydeman, Emma S Chambers, Weigang Cai, Wenhao Li, Joseph M Gibbons, Corinna Pade, Áine McKnight, Seif O Shaheen, Alex G Richter, Adrian R Martineau

## Abstract

In this population-based cohort of 7530 adults, combined IgG/A/M anti-Spike titres measured after SARS-CoV-2 vaccination were predictive of protection against breakthrough SARS-CoV-2 infection. Discrimination was significantly improved by adjustment for factors influencing risk of SARS-CoV-2 exposure including household overcrowding, public transport use, and visits to indoor public places.

## Introduction

Identifying a robust correlate of vaccine-induced protection against SARS-CoV-2 could provide insights into mechanisms of protective immune responses and accelerate licensure for candidate SARS-CoV-2 vaccines.^1^ Neutralising antibody titres have been shown to correlate with vaccine efficacy and risk of post-vaccination SARS-CoV-2 infection,^2,3^ but their determination is both resource and time intensive. Existing studies evaluating neutralising antibody titres as potential correlates of vaccine-induced protection have largely been conducted in clinical trial populations,^2,4,5^ thereby limiting generalisability of their findings to the general population. Moreover, such trials do not routinely collect data on factors influencing post-vaccination SARS-CoV-2 exposure, which precludes adjustment for these factors in statistical analyses.

Here, we explore the predictive strength of a new marker—combined IgG, IgA, and IgM responses to the SARS-CoV-2 trimeric spike glycoprotein (anti-Spike IgG/A/M)—which can be assayed in dried blood spots at low cost. We hypothesised that anti-Spike IgG/A/M titres would correlate positively with authentic virus neutralising antibody titres, and that higher titres would therefore associate with reduced risk of breakthrough infection in vaccinated individuals.

## Methods

COVIDENCE UK is a prospective, longitudinal, population-based observational study of COVID-19 in the UK general population, launched on May 1, 2020 (https://www.qmul.ac.uk/covidence). Inclusion criteria were age 16 years or older and UK residence at enrolment, with no exclusion criteria. Participants were asked to complete an online baseline questionnaire and monthly follow-up questionnaires to capture information on SARS-CoV-2 vaccine status, incident SARS-CoV-2 infection, and a wide range of potential determinants of vaccine response and SARS-CoV-2 exposure. Further details on COVIDENCE UK have been published elsewhere.^6,7^ COVIDENCE UK is registered with ClinicalTrials.gov, NCT04330599.

After enrolment into COVIDENCE UK, participants were invited to take part in a post-vaccination antibody study. The additional inclusion criterion for this study was having received a full primary course of SARS-CoV-2 vaccination (ie, two vaccine doses, or one dose of a one-dose regimen). Fully vaccinated participants were sent a kit containing instructions, lancets, and blood spot collection cards, which were returned to the study team and analysed at the Clinical Immunology Service, Institute of Immunology and Immunotherapy of the University of Birmingham (Birmingham, UK). Semi-quantitative determination of antibody titres in dried blood spot eluates was done using a commercially available ELISA that detects anti-Spike IgG/A/M with 98.3% specificity and 98.6% sensitivity (product code MK654; The Binding Site, Birmingham, UK). Further details on the assay have been published elsewhere.^7-9^ A subset of study participants attended an in-person visit, providing blood samples that permitted the measurement of neutralising antibody titres in serum using an authentic virus (Wuhan Hu-1 strain) neutralisation assay^10^ and determination of IFN-γ concentrations in supernatants of whole blood stimulated for 24 h with a pool of lyophilised peptides covering the complete protein coding sequence of the SARS-CoV-2 S glycoprotein (Miltenyi Biotech; final concentration 0.6 nM).^10^ Correlations between these responses were evaluated using Spearman ‘s rank tests.

Statistical analysis of breakthrough SARS-CoV-2 infection included data from participants in the antibody study who had not received a third or booster dose, and for whom a post-vaccination anti-Spike IgG/A/M titre was available. Participants were followed up from the day of their blood sample up to 13 days after their third or booster vaccination (if received). Breakthrough infections were defined as a reported positive result on an RT-PCR or lateral flow test for SARS-CoV-2. As duration of follow-up is a strong predictor of breakthrough infection, we truncated each participant ‘s follow-up at 18 weeks; this duration was chosen to maximise both the number of participants and the number of breakthrough infections included in the analysis. We used receiver operating characteristic (ROC) curve analysis to assess the strength of anti-Spike IgG/A/M titres as a predictor of breakthrough infection in all participants, and then stratified by vaccine type. We first included anti-Spike IgG/A/M titres as a continuous predictor in a minimally adjusted logistic regression model, adjusting for weeks between the date of full vaccination and provision of the blood sample. We then further adjusted for variables reflecting risk of SARS-CoV-2 exposure, comprising household factors (sharing a home with schoolchildren or working-age adults, and number of people per bedroom) and monthly behaviours recorded up to breakthrough infection or the end of follow-up (travel outside of the UK and number of weekly visits to the shops, to other indoor public spaces, on public transport, or to or from other households). Finally, we compared the area under the ROC curve between the minimally and fully adjusted models using Stata ‘s roccomp command. Statistical analyses were done using Stata/MP (version 17.0) and GraphPad Prism (version 9.1.2).

## Results

113 fully vaccinated participants with data on anti-Spike IgG/A/M, neutralising antibodies, and Spike peptide-stimulated IFN-γ were included in the correlation analysis, of whom 71 (62.8%) were female and 107 (94.7%) were White. 79 (69.9%) participants received a full course of ChAdOx1 n-CoV-19 (Oxford-AstraZeneca; hereafter ChAdOx1) and 34 (30.1%) a full course of BNT162b2 (Pfizer-BioNTech). Samples were taken a median of 57 days (IQR 47–71) after completion of a full primary vaccination course. Correlation was high between anti-Spike IgG/A/M and neutralising antibody titres (*r* = 0.80), but low between anti-Spike IgG/A/M titres and S peptide-stimulated IFN-γ concentrations (*r* = 0.31; figure A, B). Similar results were observed when this analysis was stratified by vaccine type (ChAdOx1: *r* = 0.71 [95% CI 0.56– 0.81], p<0.0001, and *r* = 0.35 [95% CI 0.13–0.53], p=0.0018, for anti-Spike IgG/A/M and IFN-γ, respectively; BNT162b2: *r* = 0.81 [95% CI 0.65–0.91], p<0.0001, and *r* = 0.38 [95% CI 0.04– 0.64], p=0.26, respectively).

**Figure:**
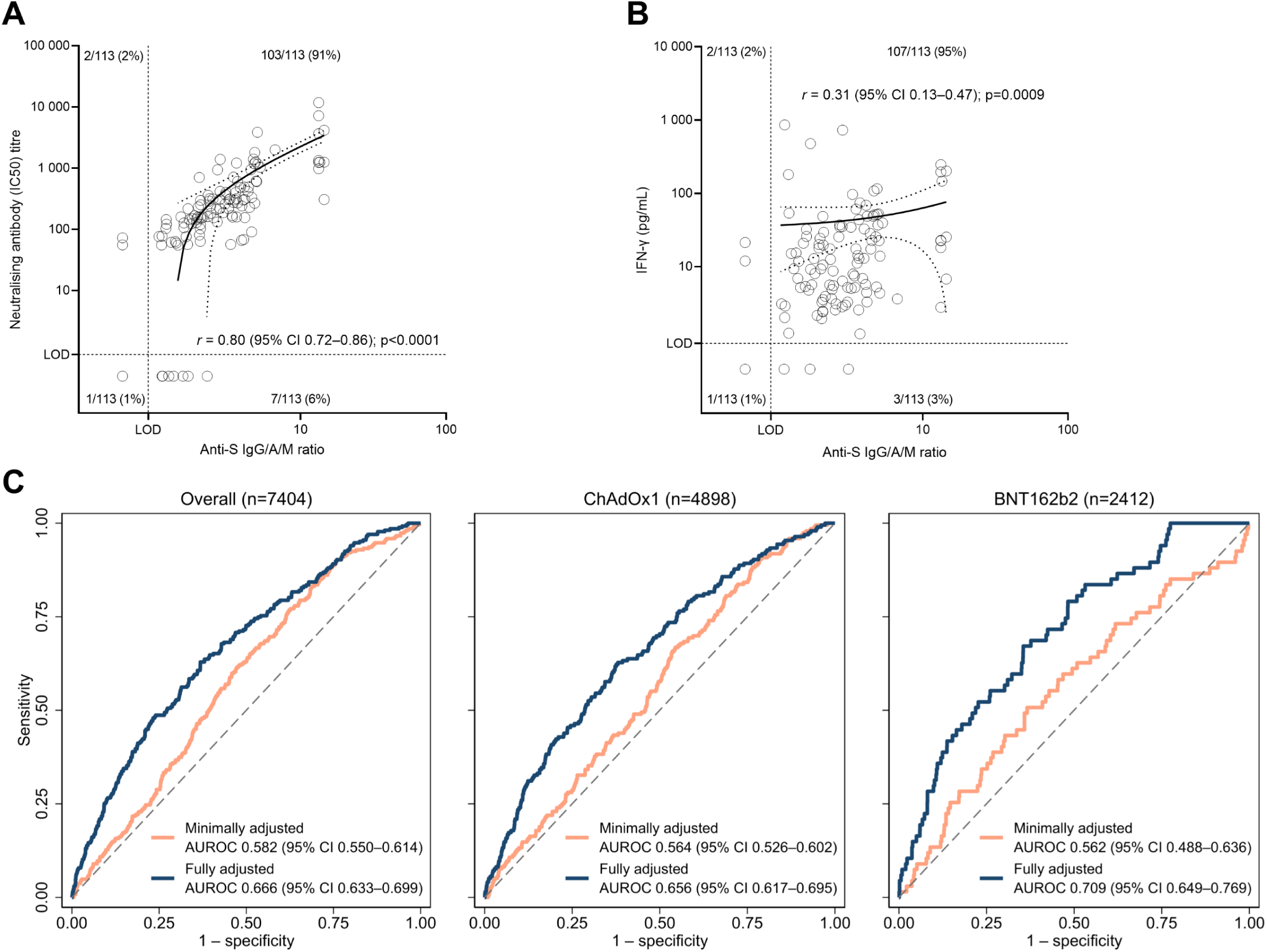
Correlation between anti-Spike IgG/A/M and neutralising antibody (IC50) titres (A) and anti-Spike IgG/A/M and IFN-γ (B), and ROC curve analysis for breakthrough SARS-CoV-2 infection after SARS-CoV-2 vaccination. (A, B) Correlation analyses were done in 113 fully vaccinated participants; distributions are shown with linear regression lines and 95% CIs. (C) ROC curves and AUROC for anti-Spike IgG/A/M ratios as a predictor of breakthrough infection, alone or adjusted for variables reflecting SARS-CoV-2 exposure, for all participants with data for included covariates and by vaccine type. IC50 = half maximal inhibitory concentration. Anti-Spike IgG/A/M = combined anti-Spike IgG, IgA, and IgM responses. IFN-γ = interferon-γ. LOD = limit of detection. AUROC = area under the receiver operating characteristic curve. ChAdOx1 = ChadOx1 nCoV-19.

7530 fully vaccinated participants, 5358 (71.2%) of whom were female, were included in our breakthrough infection analysis, with a median age of 64.2 years (IQR 57.5–69.7). 7293 (96.9%) participants were White, 71 (0.9%) were Asian, and 22 (0.3%) were Black, with the remaining 144 (1.9%) being of mixed or other ethnic groups. 367 (4.9%) participants were immunocompromised. The distribution of baseline characteristics was similar to those reported previously for the whole COVIDENCE UK cohort.^7^ 4979 (66.1%) participants received two doses of ChAdOx1 and 2455 (32.6%) participants received two doses of BNT162b2, with most remaining participants either receiving NVX-CoV2373 (Novavax; 37 [0.5%] participants) or a combination of ChAdOx1 and BNT162b2 (31 [0.4%] participants). Blood samples were provided a median of 56 days (IQR 43–68) after full vaccination, and follow-up ended a median of 26 weeks (24–28) after full vaccination.

Between Jan 12, 2021, and Jan 31, 2022, 300 (4.0%) participants reported a breakthrough SARS-CoV-2 infection during their 18 weeks of follow-up (220 [4.4%] ChAdOx1 recipients and 75 [3.1%] BNT162b1 recipients), at a median of 83 days (IQR 50–110) after their serology sample and 139 days (107–166) after full vaccination. Most infections were mild, with 28 (9%) being asymptomatic and seven (2%) requiring hospitalisation. In minimally adjusted models, anti-Spike IgG/A/M ratios were modestly predictive of breakthrough infection, but not significantly so among BNT162b2 recipients (figure C). Adjustment for factors influencing SARS-CoV-2 exposure significantly improved discrimination of the models overall (p<0.0001) and by vaccine type (ChAdOx1: p<0.0001; BNT162b1: p=0.0012; figure C).

## Discussion

In this analysis of a large population-based study, we show a strong correlation between anti-Spike IgG/A/M and neutralising antibody titres, and a far weaker correlation between anti-Spike IgG/A/M titres and S peptide-stimulated whole blood IFN-γ responses. Anti-Spike IgG/A/M titres were modestly predictive of breakthrough SARS-CoV-2 infection in the first 6– 7 months after full vaccination, with discrimination significantly improved by including key exposure variables, such as sharing a home with schoolchildren or working-age adults.

The modest discrimination of anti-Spike IgG/A/M titres for predicting breakthrough SARS-CoV-2 infection casts into doubt their effectiveness as a standalone correlate of protection. Previous studies have shown strong correlations between post-vaccination antibody titres and vaccine efficacy,^2,5^ and some have estimated anti-Spike IgG thresholds of protection for different vaccines.^4,5,11^ However, these thresholds can differ by the mechanism generating the antibodies,^11^ and may be less accurate for mild or asymptomatic infections.^4^ Additionally, few studies have considered post-vaccination exposures^11^ or explored how they work alongside antibody markers to influence risk.

The high correlation observed between neutralising antibody and anti-Spike IgG/A/M titres confirms and extends previous findings of correlation with anti-Spike IgG antibodies^4,5^ by additionally including IgA and IgM responses. However, the modest predictive ability of anti-Spike IgG/A/M suggests that it fails to capture a key element of protection, such as cellular responses. This is supported by the weak correlation we observed between anti-Spike IgG/A/M titres and S peptide-stimulated IFN-γ concentrations. Although humoral and cellular responses are sometimes correlated, they can be discordant,^12^ suggesting that anti-Spike IgG/A/M alone will not be sufficient to predict risk of breakthrough infection after SARS-CoV-2 vaccination.

Among the strengths of this analysis is the population-based nature of the cohort. Previous studies on correlates of protection have largely relied on post-hoc analyses of vaccine trials,^2,4,5^ featuring a wider range of inclusion and exclusion criteria. Our cohort therefore provides a more general and diverse population, including participants with poor health, various comorbidities, and immunosuppression. Additionally, our monthly questionnaires allow a detailed evaluation of exposure risks, capturing time-varying behaviours such as interactions with other households. Finally, we used a sensitive CE-marked, validated assay that extends beyond IgG and captures combined antibody responses.

A limitation of our study is that our findings are limited to recipients of ChAdOx1 and BNT162b2 vaccines, although these are two of the most widely administered vaccines and operate via different mechanisms. We also lacked statistical power in our analysis of breakthrough infection following vaccination with BNT162b2, because of fewer participants and lower rates of infection. Finally, our results are not generalisable to children or populations of non-White ethnicity.

In conclusion, our combined measure of anti-Spike IgG/A/M was only modestly predictive of breakthrough infection in vaccinated individuals, and performance was significantly improved by incorporating factors reflecting post-vaccination SARS-CoV-2 exposure. Combined with the poor correlation observed between anti-Spike IgG/A/M and S peptide-stimulated IFN-γ, these findings suggest that the relatively limited predictive value of anti-Spike IgG/A/M titres for protection may reflect their inability to reflect protective vaccine-induced cellular responses.

## Data Availability

De-identified participant data will be made available upon reasonable request to the corresponding author.

## Contributors

ARM wrote the study protocol, with input from HH, MT, and SOS. HH, MT, SF, and ARM contributed to questionnaire development and design. HH co-ordinated and managed the study, with input from ARM, DAJ, MT, SOS, and NP. HH, ARM, and SOS supported recruitment. SF and AGR developed, validated, and performed assays for Anti-S IgG/A/M antibodies. EC, WC and WL performed S peptide-stimulated whole blood assays. JG, CP and AM developed, validated, and performed assays for neutralising antibodies. GV, DAJ, FT, MT, and HH contributed to data management. GV did the statistical analysis, with input from ARM. GV and ARM wrote the first draft of the report. All authors revised it critically for important intellectual content, gave final approval of the version to be published, and agreed to be accountable for all aspects of the work in ensuring that questions related to the accuracy or integrity of any part of the work were appropriately investigated and resolved. ARM had final responsibility for the decision to submit for publication.

## Declaration of interests

ARM declares receipt of funding in the last 36 months to support vitamin D research from the following companies who manufacture or sell vitamin D supplements: Pharma Nord, DSM Nutritional Products, Thornton & Ross, and Hyphens Pharma. ARM also declares support for attending meetings from the following companies who manufacture or sell vitamin D supplements: Pharma Nord and Abiogen Pharma. ARM also declares participation on the Data and Safety Monitoring Board for the Chair, DSMB, VITALITY trial (Vitamin D for Adolescents with HIV to reduce musculoskeletal morbidity and immunopathology). ARM also declares unpaid work as a Programme Committee member for the Vitamin D Workshop. ARM also declares receipt of vitamin D capsules for clinical trial use from Pharma Nord, Synergy Biologics, and Cytoplan.

## Funding

This study was supported by a grant from Barts Charity to ARM (MGU0466), and by donations to Queen Mary University of London by the Fischer Family Trust, The Exilarch ‘s Foundation, and DSM Nutritional Products. DAJ and ESC are supported by a grant from Barts Charity (MGU0459). MT is supported by a grant from the Rosetrees Trust and The Bloom Foundation (M771). AMK and JMG are supported by grants from the Rosetrees Trust (CF1\100003) and Barts Charity (MGU0558). AMK is also supported by the John Black Charitable Foundation (M946). The views expressed are those of the authors and not necessarily those of the funders.

## Notes

### Author Declarations

COVIDENCE UK was sponsored by Queen Mary University of London and approved by Leicester South Research Ethics Committee (ref 20/EM/0117).

